# Prevalence and Correlates of Mild Cognitive Impairment Among Older Adults in Ho Chi Minh City: A Cross-Sectional Study

**DOI:** 10.1101/2025.10.09.25337695

**Authors:** Khai Quang Nguyen, The Ha Ngoc Than, Kien Gia To

## Abstract

**Introduction:** This study aims to estimate the prevalence of mild cognitive impairment (MCI) and identify associated sociodemographic, lifestyle, and functional factors among older adults attending outpatient care in a secondary-level clinical setting in Vietnam.

**Methods:** A cross-sectional study was conducted from May 2024 to February 2025. Participants were adults aged 60 years or older, literate in Vietnamese, and able to complete the cognitive screening protocol. Data were collected through structured interviews, medical record reviews, and assessments using the Mini-Mental State Examination (MMSE) and the Lawton Instrumental Activities of Daily Living (IADL) scale.

**Results:** Of the 631 participants, 30.1% screened positive for MCI. Factors significantly associated with MCI included older age, being unmarried, unstable income, lower educational attainment, reduced recreational activity, and lower IADL scores. Multivariable logistic regression analysis confirmed these associations, with older age and reduced recreational activity showing the strongest links to MCI.

**Conclusion:** The high prevalence of MCI among older outpatients highlights the need for integrating cognitive and social-functional screening into outpatient care. Addressing social vulnerabilities and promoting recreational activities may help mitigate cognitive decline in this population.

## I. INTRODUCTION

Vietnam is undergoing a rapid demographic transition, with the proportion of individuals aged 65 and older projected to double from 7% in 2020 to 14% by 2035, a rate faster than in many developed countries [1]. By 2024, adults aged 60 and above already accounted for over 14% of the population [2]. This aging shift brings growing health challenges, notably cognitive decline. Modeling studies predict a sharp rise in cognitive impairment across Southeast Asia, including Vietnam [3].

Mild cognitive impairment (MCI) is a transitional state between normal ageing and dementia [4]. Although basic daily functioning is preserved, MCI affects higher-order cognitive and social abilities [5]. Its burden extends to economic and survival consequences, with annual care costs exceeding 24,000 USD in the United States [6] and high five-year mortality reported among older adults with dementia in low- and middle-income countries (LMICs) [7].

In Vietnam, many older adults bypass lower-level facilities and seek care at secondary-level hospitals [8]. Memory and cognitive concerns are increasingly observed in this outpatient population. Most studies on MCI in Vietnam and neighbouring countries have used community samples, potentially overlooking this high-risk group [9-12]. Information routinely collected at triage, including social and functional data, may provide a practical opportunity for early recognition [13].

Given the demographic pace of aging and the strain on hospital services in Vietnam [14], understanding the prevalence and correlates of MCI in outpatient care is essential for developing sustainable models of geriatric care. We therefore conducted a hospital-based study to estimate the prevalence of screen-detected MCI and to identify associated sociodemographic, lifestyle, and functional factors among older adults attending outpatient care in a secondary-level clinical setting in Vietnam.

## II. METHODS

### Study design and participants

A cross-sectional study was conducted among older adults attending the outpatient department of a secondary-level public hospital in southern Vietnam between May 2024 and February 2025. Ethical approval was obtained from relevant institutional review boards, and written informed consent was obtained from all participants.

Eligible participants were adults aged 60 years or older, literate in Vietnamese, and able to complete the full cognitive screening protocol. Exclusion criteria included uncorrected sensory impairments, acute medical conditions affecting attention, psychiatric disorders unrelated to dementia, significant depressive symptoms (GDS-15≥5), current use of medications with potential cognitive side effects, and recent participation in structured cognitive training programs.

Screening began with a brief conversation to confirm willingness to participate and language fluency before applying formal eligibility criteria. Individuals who sought only to check their MMSE score, declined written consent, or withdrew after partial participation were excluded.

### Data collection and tools

Before recruitment, the research team coordinated with outpatient administrative staff to identify optimal time periods when older adults most frequently attended consultations. This minimized disruption to clinical operations and facilitated natural engagement with potential participants in waiting areas.

To reduce reluctance in discussing memory concerns, interviews began with general health topics before cognitive questions. All assessments were conducted by trained researchers. Medication use and chronic conditions were verified through medical-record review and consultation with treating physicians. Demographic data including age, gender, marital status, education, and financial security were obtained through structured face-to-face interviews.

#### Recreational activities

Social recreational engagement was assessed through interviewer-administered questions about recent participation in activities that involved interaction with other people. Activities undertaken alone were not considered. Participants reporting such engagement were asked about frequency, and their participation was categorized as regular when occurring at least three times per week, each lasting ≥30 minutes and sustained for at least one month. Less frequent participation was considered irregular, while those reporting none were classified as having no engagement.

#### Hypertension and diabetes status

Hypertension and diabetes mellitus were each categorized as no documented diagnosis, controlled, or uncontrolled, based on medical record review and participant self-report. No documented diagnosis indicated absence of both patient-reported history and chart documentation. Controlled referred to a documented condition with stable treatment, no recent medication change or hospitalization (within three months), and recent clinical values within target ranges (BP <140/90 mmHg; HbA1c <7.5%).

#### Physical activity-limiting conditions

Defined as any acute or chronic condition that significantly restricted mobility or posed exertional risk. Verification included both self-report and review of medical instructions, supplemented by contextual questions (“Has any doctor told you to avoid certain exercises?”).

#### Functional status

Functional autonomy was evaluated using the culturally adapted Vietnamese version of the Lawton Instrumental Activities of Daily Living (IADL) scale [15]. The scale evaluates eight domains, including use of the telephone, shopping, food preparation, housekeeping, laundry, transportation, medication management, and financial handling. Scores range from 0 to 8, with higher scores indicating greater functional autonomy.

### Criteria for MCI Classification

MCI in this study was identified based on (i) self-reported memory complaints, (ii) Mini-Mental State Examination (MMSE) scores of 24-29, (iii) relatively preserved instrumental functioning on the Lawton IADL scale (score ≥5) [16], and (iv) absence of a clinical diagnosis of dementia [4,17]. The Vietnamese version of the MMSE, officially endorsed by the Ministry of Health, was used to ensure linguistic and cultural validity.

The MMSE range of 24–29 was selected based on field considerations and published evidence indicating that scores below 24 often reflect dementia, whereas higher values may still capture subtle impairments [18-20]. This range was appropriate for outpatient screening contexts in low-resource settings.

The MMSE and cut-off values were employed to identify early cognitive changes consistent with MCI risk, rather than to establish definitive diagnoses. Hence, prevalence estimates represent individuals at potential risk rather than confirmed MCI cases, favoring sensitivity over specificity. To confirm the absence of diagnosed dementia, medical records were reviewed. Participants with borderline MMSE scores (24), inconsistent interview responses, or caregiver-reported cognitive concerns were referred for further evaluation by an independent neurologist or geriatrician.

### Pilot study

A preliminary pilot study tested the feasibility and reliability of the screening protocol and instruments (MMSE, GDS-15, Lawton IADL) before full-scale implementation [21]. The pilot achieved an 89.4% enrollment rate, 98% screening completion, and full adherence among eligible participants. Internal consistency was acceptable: MMSE (α=0.72), GDS-15 (α=0.71), and Lawton IADL (α=0.73), supporting their use in this main study.

### Statistical methods

All analyses were performed using SPSS version 27. Descriptive statistics summarized sample characteristics. Group comparisons between participants with and without MCI used independent t-tests or Mann–Whitney U tests for continuous variables, and chi-square or Fisher’s exact tests for categorical variables.

Univariate logistic regressions were first performed for all candidate factors. Variables with p<0.25 were entered into the multivariable model to avoid excluding potentially relevant factors [22]. For categorical variables with more than two levels, overall significance was assessed.

Multicollinearity was assessed using variance inflation factors (VIF), with values >2.5 indicating potential collinearity [23]. Model calibration was evaluated with the Hosmer– Lemeshow goodness-of-fit test to verify alignment between predicted probabilities and observed MCI frequencies across risk deciles [24]. A non-significant result indicated acceptable calibration. All statistical tests were two-tailed, and significance was defined as p<0.05.

## III. RESULTS

### 3.1. Characteristics of the participants

Of the 726 older adults approached, 653 (89.9%) agreed to screening. Twenty-two individuals were excluded: seven preferred to complete only the MMSE, seven declined to complete the MMSE, three scored ≥5 on the GDS-15, three had uncorrected sensory impairments, and two were taking medications with cognitive side effects. The final analytical sample comprised 631 participants (*Figure 1*).

**FIGURE 1.**
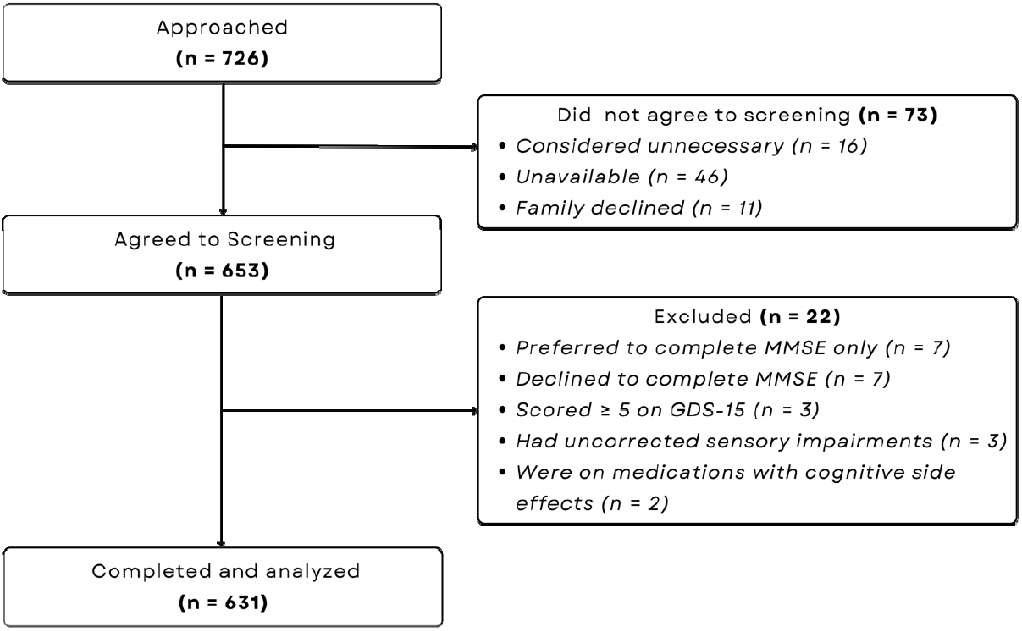
Flow chart of the study.

Among the 631 participants (*Table 1*), the average age was 65.9 years (SD=5.3), and 57.7% were female. Most were married or cohabiting (56.3%), had completed senior high school (75.3%), and reported financial stability (72.1%). Recreational engagement varied: 30.6% reported no participation, and only 22.6% met criteria for regular involvement. The average Lawton IADL score was 6.8 (SD=0.9), and physical limitations were rare (1.4%). Regarding metabolic health, 46.7% had diabetes, of whom 27.0% were uncontrolled; hypertension was present in 63.8%, with 13.4% uncontrolled.

**TABLE 1.** Demographic characteristics of participants.

| **Variables** | **Total**<br>**(n=631)** | **MCI**<br>**(n = 190)** | **Non-MCI**<br>**(n = 441)** | **p** |
| --- | --- | --- | --- | --- |
| Age (Mean ± SD) | 65.91 ± 5.31 | 70.19 ± 4.92 | 64.06 ± 4.33 | <0.001 |
| Gender (%) |  |  |  | 0.099 |
| Female | 364 (57.7) | 119 (62.6) | 245 (55.6) |  |
| Male | 267 (42.3) | 71 (37.4) | 196 (44.4) |  |
| Marital status (%) |  |  |  | <0.001 |
| Single/ Divorced/ Separated/<br>Widowed | 276 (43.7) | 123 (64.7) | 153 (34.7) |  |
| Married or Cohabiting | 355 (56.3) | 67 (35.3) | 288 (65.3) |  |
| Financial security (%) |  |  |  | <0.001 |
| Unstable income | 176 (27.9) | 78 (41.1) | 98 (22.2) |  |
| Stable income | 455 (72.1) | 112 (58.9) | 343 (77.8) |  |
| Education (%) |  |  |  | <0.001 |
| Less than senior high school | 156 (24.7) | 82 (43.2) | 74 (16.8) |  |
| Completed senior high school | 475 (75.3) | 108 (56.8) | 367 (83.2) |  |
| Recreational activities (%) |  |  |  | <0.001 |
| None | 193 (30.6) | 107 (56.3) | 86 (19.5) |  |
| Irregularity | 295 (46.8) | 73 (38.4) | 222 (50.3) |  |
| Regularity | 143 (22.6) | 10 (5.3) | 133 (30.2) |  |
| HTN Status (%) |  |  |  | 0.185 |
| No documented diagnosis | 229 (36.2) | 73 (38.4) | 156 (35.4) |  |
| Controlled | 348 (55.2) | 96 (50.5) | 252 (57.1) |  |
| Uncontrolled | 54 (8.6) | 21 (11.1) | 33 (7.5) |  |
| DM Status (%) |  |  |  | 0.001 |
| No documented diagnosis | 336 (53.3) | 83 (43.7) | 253 (57.4) |  |
| Controlled | 216 (34.2) | 85 (44.7) | 131 (29.7) |  |
| Uncontrolled | 79 (12.5) | 22 (11.6) | 57 (12.9) |  |
| Condition limits or contraindicates PA (%) |  |  |  | >0.999* |
| No | 622 (98.6) | 187 (98.4) | 435 (98.6) |  |
| Yes | 9 (1.4) | 3 (1.6) | 6 (1.4) |  |
| Lawton IADL (Mean ± SD) | 6.81 ± 0.85 | 6.22 ± 0.69 | 7.07 ± 0.78 | <0.001 |
| **Abbreviations:** |  |  |  |  |
| MCI, Mild Cognitive Impairment; DM, Diabetes Mellitus; HTN, Hypertension; PA, Physical Activity; Lawton IADL, Lawton Instrumental Activities of Daily Living Scale; SD, Standard Deviation. |  |  |  |  |
| **Footnotes:** |  |  |  |  |
| The continuous variables were shown as mean ± SD; the categorical variables were shown as percentage. | | | | |
| p-values were calculated using Chi-square test or Fisher's Exact test (*) for categorical variables and independent-sample t-test for continuous variables. |  |  |  |  |

MCI was identified in 190 of 631 participants, yielding a prevalence of 30.1% (95%CI: 26.5%–33.7%), based on fulfillment of four criteria [4,17]: (i) self-reported memory complaints, (ii) MMSE 24-29, (iii) Lawton IADL ≥5 [16], and (iv) absence of a dementia diagnosis. Overall, 61.3% reported subjective memory complaints, 30.1% scored within the MMSE range, and all met functional and diagnostic exclusion criteria.

Compared to non-MCI individuals, those with MCI were older (mean age 70.2 vs. 64.1 years, p<0.001), more often not partnered (64.7% vs. 34.7%, p<0.001), less likely to have completed senior high school (56.8% vs. 83.2%, p<0.001), and more often reported unstable income (41.1% vs. 22.2%, p<0.001). Gender distribution did not significantly differ (p=0.099).

Recreational inactivity was markedly higher among the MCI group (56.3% vs. 19.5%, p<0.001), whereas regular participation was lower (5.3 % vs. 30.2 %). Functional scores were reduced (mean Lawton =6.22 vs. 7.07, p<0.001). Diabetes status differed by group (p=0.001), whereas hypertension (p=0.185) and physical activity-limiting conditions (p>0.999) showed no significant difference.

### 3.2. Univariate and multivariate analysis

Univariate logistic regression identified older age, unmarried status, unstable income, lower education, reduced recreational and IADLs activity as factors associated with higher MCI odds (all p<0.05). Variables with p<0.25 were included in the multivariable model [22]; hypertension and diabetes did not meet this criterion (*Table 2*).

**TABLE 2.**
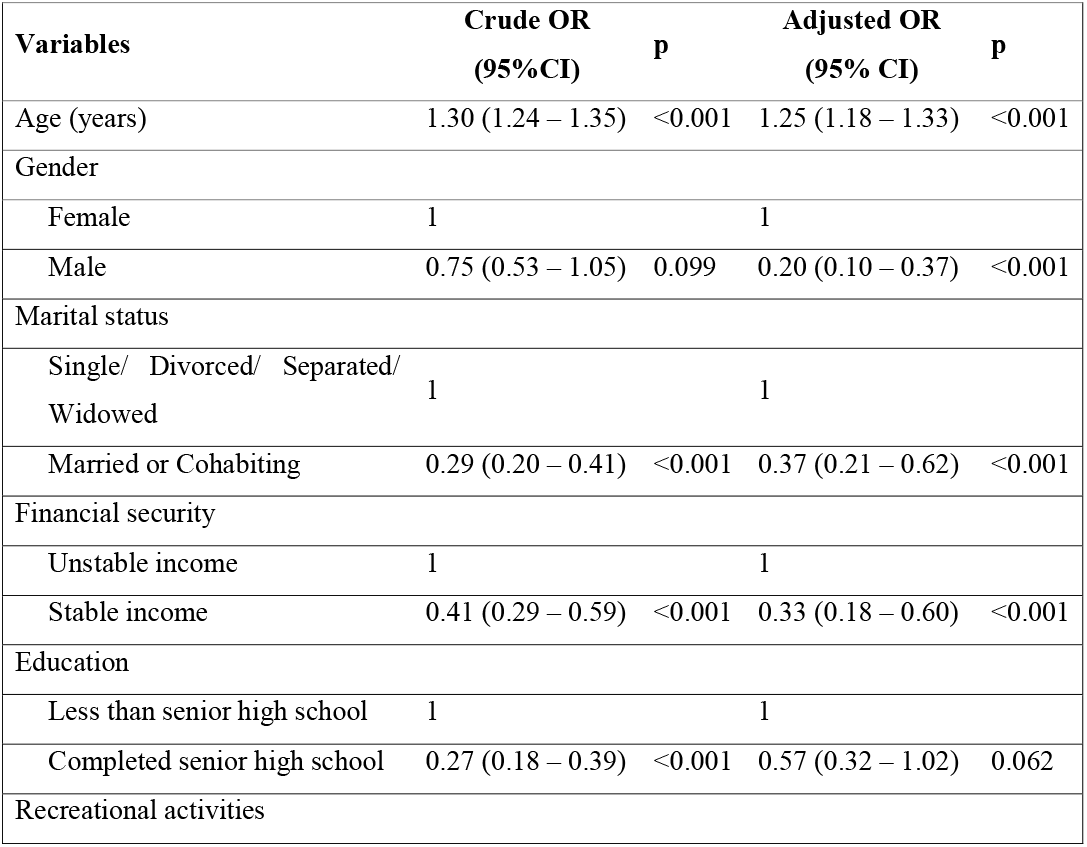
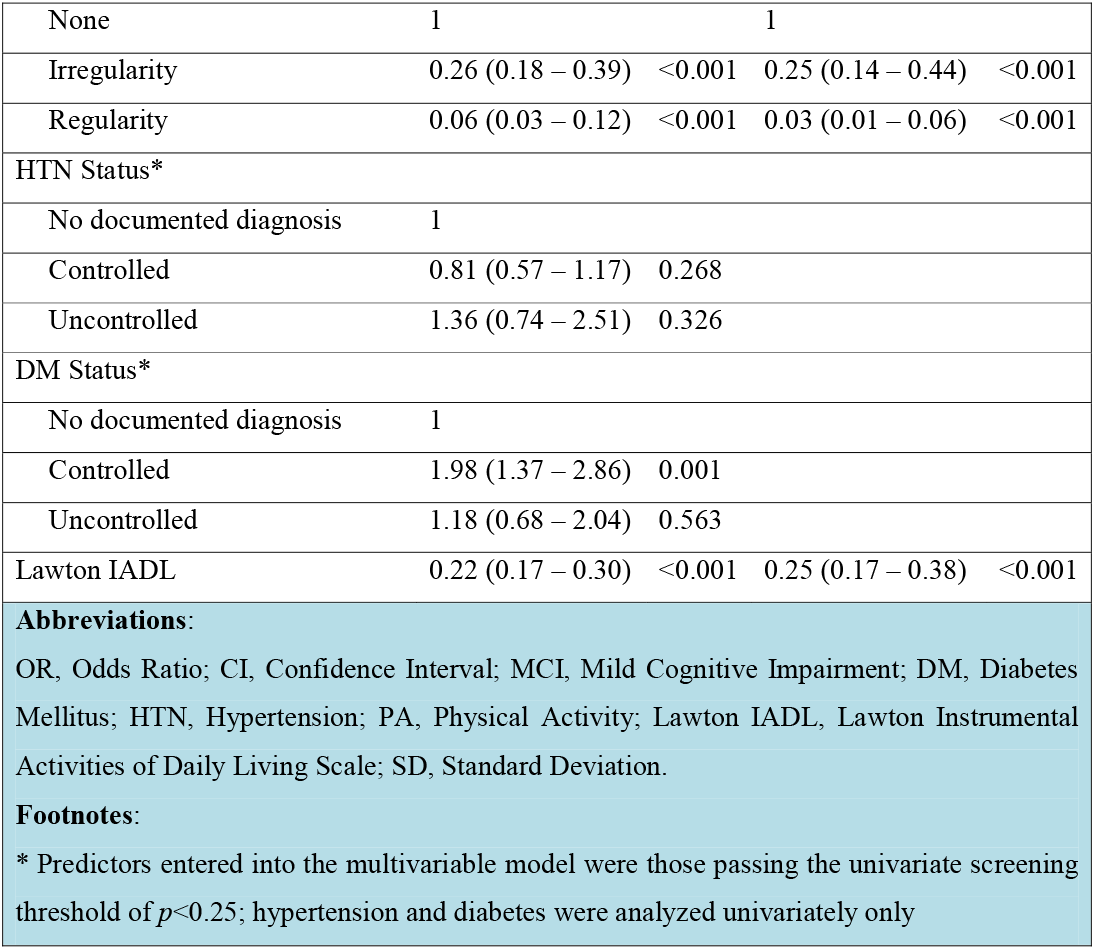
Univariate and multivariate logistic regression analysis of factors associated with MCI.

In the adjusted model, older age remained strongly associated with MCI (aOR=1.25; p<0.001). Male sex (aOR=0.20, p<0.001), being married/cohabiting (aOR=0.37, p<0.001), and stable income (aOR=0.33, p<0.001) were protective. Education showed a borderline association (aOR=0.57; p=0.062). Recreational activity was inversely associated with MCI, with irregular (aOR=0.25, p<0.001) and regular participation (aOR=0.03, p<0.001) linked to lower odds compared with no activity. Higher IADL scores remained independently protective (aOR=0.25, p<0.001). No multicollinearity was detected (all VIFs<1.5), and model fit was acceptable (Hosmer–Lemeshow χ^2^=10.46, p=0.234).

## IV. DISCUSSION

This study found that nearly one in three older adults attending a secondary-level outpatient department screened positive for MCI. Risk was associated with age, gender, marital status, financial security, recreational activity, and functional capacity. Such findings confirm that cognitive vulnerability is common in clinical settings and closely interwoven with social context.

The observed prevalence aligns with the upper range of estimates reported across Asia. A Thai urban community study using the Montreal Cognitive Assessment reported 33.9% prevalence among adults aged 30–65 years [12], while a Myanmar study using the Revised Hasegawa’s Dementia Scale found 29.9% among those aged ≥60 [9]. In contrast, a pooled Chinese meta-analysis of 41 community-based studies reported a lower prevalence of 12.2%, largely reflecting variations in diagnostic tools and age composition [25]. A Malaysian longitudinal study documented an incidence rate of 10.5 per 100 person-years among cognitively intact older adults over 1.5 years [10]. Such variation reflects methodological and population differences. The hospital-based context likely includes more individuals concerned about cognitive change, distinguishing them from general community samples (28.2%) reported in Hue City, Vietnam [11].

Older age was the strongest factor associated with impairment, with affected participants being on average six years older than those without impairment. This aligns with neurobiological evidence linking advancing age to cognitive decline [26]. In Vietnam, demographic shifts are amplifying this concern. A large cross-sectional study of 3,500 older adults reported a sharp rise in cognitive symptoms after age 70 [27], supporting the need for age-sensitive cognitive screening in geriatric care.

Social circumstances also influenced risk. Being married or cohabiting was protective, consistent with a Vietnamese cohort showing that individuals aged ≥60 living with both spouse and children had better cognitive performance than those living alone, suggesting a potential protective role of co-residence and familial support [28].

Conversely, participants reporting unstable income had nearly threefold higher odds of impairment. National surveys indicate that financial strain among older households in Vietnam contributes to delayed medical visits and reduced care access [29]. International evidence also links financial hardship to poorer diet quality and fewer cognitively stimulating activities, potentially reducing cognitive reserve [30].

Lifestyle engagement also differentiated cognitive risk. A dose–response pattern was observed, as even occasional recreational activity was associated with lower odds of impairment, while regular participation provided the greatest protection. This aligns with evidence that socially and cognitively stimulating activities enhance neural plasticity and slow decline [31]. Longitudinal data further indicate a bi-directional relationship, as participation both preserves cognition and is sustained by better cognitive function, forming a reinforcing cycle of resilience [32].

Functional limitations in IADLs were closely linked to MCI. Lower Lawton scores among affected individuals suggest that early deficits in managing complex daily tasks may signal progression toward dementia [33]. Yet, IADL performance is influenced by other domains such as mobility, balance, and sensory function [34]. Therefore, clinicians should interpret functional limitations within a multidimensional context.

Gender differences were apparent only after multivariable adjustment, with women showing greater risk of impairment. This pattern mirrors previous evidence and may reflect both biological and social factors [35,36]. Biologically, faster hippocampal atrophy, greater amyloid-tau burden, and postmenopausal estrogen decline may heighten female vulnerability [35]. Socially, lower educational and occupational complexity among older women may compound risk [36]. These interpretations warrant caution. We did not measure reporting propensity or detailed life □ course exposures [37], and the male–female health □ survival paradox may further complicate interpretation [38].

This study has limitations. The cross-sectional design precludes causal inferences. Although MMSE cut-offs were based on prior validation studies [18-20], cultural and educational differences may still influence performance [39]. Participation bias is possible because some eligible older adults declined full assessment, and the single-center, urban sample may limit generalizability, particularly to rural areas.

## V. CONCLUSION

This hospital-based study found that mild cognitive impairment affected nearly one-third of older outpatients and was linked to age, gender, social vulnerability, recreational inactivity, and functional decline. These results highlight the value of integrating brief cognitive and social-functional screening into outpatient workflows to inform more comprehensive geriatric care in Vietnam.

## Data Availability

All data produced in the present study are available upon reasonable request to the authors

## REFERENCES

1. Maheshwari A, Maheshwari G. Aging Population in Vietnam: Challenges, Implications, and Policy Recommendations. Int J Aging. 2024/3/11 2024;2(1):e1–e1. doi:10.34172/ija.2024.e1

2. General Statistics Office of Vietnam. Key Findings of the 2024 Mid-term Population and Housing Survey (as of 0:00, April 1, 2024). Statistical Publishing House; 2024.

3. Norton S, Matthews FE, Brayne C. A commentary on studies presenting projections of the future prevalence of dementia. BMC Public Health. Jan 2 2013;13:1. doi:10.1186/1471-2458-13-1

4. Petersen RC. Mild cognitive impairment as a diagnostic entity. Journal of internal medicine. Sep 2004;256(3):183–94. doi:10.1111/j.1365-2796.2004.01388.x

5. Werner P. Mild Cognitive Impairment and Caregiver Burden: A Critical Review and Research Agenda. Public Health Reviews. 2012/11/30 2012;34(2):16. doi:10.1007/BF03391684

6. Frech FH, Li G, Juday T, et al. Economic Impact of Progression from Mild Cognitive Impairment to Alzheimer Disease in the United States. J Prev Alzheimers Dis. 2024;11(4):983–991. doi:10.14283/jpad.2024.68

7. Piovezan RD, Oliveira D, Arias N, Acosta D, Prince MJ, Ferri CP. Mortality Rates and Mortality Risk Factors in Older Adults with Dementia from Low-and Middle-Income Countries: The 10/66 Dementia Research Group Population-Based Cohort Study. Journal of Alzheimer’s disease : JAD. 2020;75(2):581–593. doi:10.3233/jad-200078

8. Nguyen TA, Giang LT. Factors Influencing the Vietnamese Older Persons in Choosing Healthcare Facilities. Health Serv Insights. 2021;14:11786329211017426. doi:10.1177/11786329211017426

9. Saw YM, Saw TN, Than TM, et al. Cognitive impairment and its risk factors among Myanmar elderly using the Revised Hasegawa’s Dementia Scale: A cross-sectional study in Nay Pyi Taw, Myanmar. PloS one. 2020;15(7):e0236656. doi:10.1371/journal.pone.0236656

10. Hussin NM, Shahar S, Yahya HM, Din NC, Singh DKA, Omar MA. Incidence and predictors of mild cognitive impairment (MCI) within a multi-ethnic Asian populace: a community-based longitudinal study. BMC Public Health. 2019/08/22 2019;19(1):1159. doi:10.1186/s12889-019-7508-4

11. Duong LDN, Hoai Thuong. Cognitive impairment and sleeping disorder among the elderly at communities in Hue city, Vietnam. Journal of Preventive Medicine. 2017;27(Suppl. 3):15–22.

12. Pinyopornpanish K, Buawangpong N, Soontornpun A, et al. A household survey of the prevalence of subjective cognitive decline and mild cognitive impairment among urban community-dwelling adults aged 30 to 65. Sci Rep. Apr 2 2024;14(1):7783. doi:10.1038/s41598-024-58150-3

13. Livingston G, Huntley J, Sommerlad A, et al. Dementia prevention, intervention, and care: 2020 report of the Lancet Commission. Lancet (London, England). Aug 8 2020;396(10248):413–446. doi:10.1016/s0140-6736(20)30367-6

14. Dang TH, Nguyen TA, Hoang Van M, Santin O, Tran OMT, Schofield P. Patient-Centered Care: Transforming the Health Care System in Vietnam With Support of Digital Health Technology. J Med Internet Res. Jun 4 2021;23(6):e24601. doi:10.2196/24601

15. Graf C. The Lawton instrumental activities of daily living scale. Am J Nurs. Apr 2008;108(4):52–62; quiz 62-3. doi:10.1097/01.Naj.0000314810.46029.74

16. Ouchi Y, Kasai M, Nakamura K, Nakatsuka M, Meguro K. Qualitative Assessment of Instrumental Activities of Daily Living in Older Persons with Very Mild Dementia: The Kurihara Project. Dement Geriatr Cogn Dis Extra. May-Aug 2016;6(2):374–381. doi:10.1159/000446769

17. Albert MS, DeKosky ST, Dickson D, et al. The diagnosis of mild cognitive impairment due to Alzheimer’s disease: recommendations from the National Institute on Aging-Alzheimer’s Association workgroups on diagnostic guidelines for Alzheimer’s disease. Alzheimer’s & dementia : the journal of the Alzheimer’s Association. May 2011;7(3):270–9. doi:10.1016/j.jalz.2011.03.008

18. Feng L, Chong MS, Lim WS, Ng TP. The Modified Mini-Mental State Examination test: normative data for Singapore Chinese older adults and its performance in detecting early cognitive impairment. Singapore Med J. Jul 2012;53(7):458–62.

19. Zhang S, Qiu Q, Qian S, et al. Determining Appropriate Screening Tools and Cutoffs for Cognitive Impairment in the Chinese Elderly. Front Psychiatry. 2021;12:773281. doi:10.3389/fpsyt.2021.773281

20. Salis F, Costaggiu D, Mandas A. Mini-Mental State Examination: Optimal Cut-Off Levels for Mild and Severe Cognitive Impairment. Geriatrics (Basel). Jan 12 2023;8(1) doi:10.3390/geriatrics8010012

21. Nguyen KQ, Than THN, To KG. Early screening for mild cognitive impairment among adults aged 60 and above: A pilot study at a secondary hospital in Ho Chi Minh City, Vietnam. medRxiv. 2025:2025.07.10.25331273. doi:10.1101/2025.07.10.25331273

22. Bursac Z, Gauss CH, Williams DK, Hosmer DW. Purposeful selection of variables in logistic regression. Source Code Biol Med. Dec 16 2008;3:17. doi:10.1186/1751-0473-3-17

23. Senaviratna NAMR, A. Cooray TMJ. Diagnosing Multicollinearity of Logistic Regression Model. Asian Journal of Probability and Statistics. 10/01 2019;5(2):1–9. doi:10.9734/ajpas/2019/v5i230132

24. Assessing the Fit of the Model. Applied Logistic Regression. 2000:143–202.

25. Lu Y, Liu C, Yu D, et al. Prevalence of mild cognitive impairment in community-dwelling Chinese populations aged over 55 □ years: a meta-analysis and systematic review. BMC Geriatr. Jan 6 2021;21(1):10. doi:10.1186/s12877-020-01948-3

26. Craik FIM, Bialystok E. Cognition through the lifespan: mechanisms of change. Trends in Cognitive Sciences. 2006/03/01/ 2006;10(3):131-138. doi:10.1016/j.tics.2006.01.007

27. Bich NN, Dung NTT, Vu T, et al. Dementia and associated factors among the elderly in Vietnam: a cross-sectional study. Int J Ment Health Syst. 2019;13:57. doi:10.1186/s13033-019-0314-7

28. Bandyopadhyay A, Korinek K, Adkins D. Cognitive Impairment In Vietnamese Older Adults: Effects Of Living Arrangements And Familial Support. Innovation in Aging. 2024;8(Supplement_1):147–148. doi:10.1093/geroni/igae098.0475

29. Giang NH, Vinh NT, Phuong HT, Thang NT, Oanh TTM. Household financial burden associated with healthcare for older people in Viet Nam: a cross-sectional survey. Health Res Policy Syst. Nov 29 2022;20(Suppl 1):112. doi:10.1186/s12961-022-00913-3

30. Shatenstein B, Ferland G, Belleville S, et al. Diet quality and cognition among older adults from the NuAge study. Exp Gerontol. May 2012;47(5):353–60. doi:10.1016/j.exger.2012.02.002

31. Zhou Y, Chen Z, Shaw I, et al. Association between social participation and cognitive function among middle- and old-aged Chinese: A fixed-effects analysis. J Glob Health. Dec 2020;10(2):020801. doi:10.7189/jogh.10.020801

32. Wang J, Li S, Hu Y, et al. The bi-directional relationships between diversified leisure activity participation and cognitive function in older adults in China: separating between-person effects from within-person effects. BMC Geriatr. May 13 2024;24(1):426. doi:10.1186/s12877-024-04997-0

33. Luck T, Luppa M, Wiese B, et al. Prediction of incident dementia: impact of impairment in instrumental activities of daily living and mild cognitive impairment-results from the German study on ageing, cognition, and dementia in primary care patients. Am J Geriatr Psychiatry. Nov 2012;20(11):943–54. doi:10.1097/JGP.0b013e31825c09bc

34. Bruderer-Hofstetter M, Gorus E, Cornelis E, Meichtry A, De Vriendt P. Influencing factors on instrumental activities of daily living functioning in people with mild cognitive disorder – a secondary investigation of cross-sectional data. BMC Geriatrics. 2022/10/11 2022;22(1):791. doi:10.1186/s12877-022-03476-8

35. Zhu D, Montagne A, Zhao Z. Alzheimer’s pathogenic mechanisms and underlying sex difference. Cell Mol Life Sci. Jun 2021;78(11):4907–4920. doi:10.1007/s00018-021-03830-w

36. Liu Y, Yu X, Han P, et al. Gender-specific prevalence and risk factors of mild cognitive impairment among older adults in Chongming, Shanghai, China. Frontiers in aging neuroscience. 2022;14:900523. doi:10.3389/fnagi.2022.900523

37. Narbutas J, Egroo MV, Chylinski D, et al. Cognitive efficiency in late midlife is linked to lifestyle characteristics and allostatic load. Aging (Albany NY). Sep 8 2019;11(17):7169–7186. doi:10.18632/aging.102243

38. Susan C. Alberts EAA, Laurence R. Gesquiere, Jeanne Altmann, James W. Vaupel, Kaare Christensen. The Male-Female Health-Survival Paradox: A Comparative Perspective on Sex Differences in Aging and Mortality. In: Weinstein M LM, ed. Sociality, Hierarchy, Health: Comparative Biodemography: A Collection of Papers. National Academies Press (US); 2014.

39. Quattropani MC, Sardella A, Morgante F, et al. Impact of Cognitive Reserve and Premorbid IQ on Cognitive and Functional Status in Older Outpatients. Brain Sci. Jun 22 2021;11(7) doi:10.3390/brainsci11070824

